# Comprehensive humoral and cellular immune responses to SARS-CoV-2 variants in diverse Chinese populations: A benefit perspective of national vaccination

**DOI:** 10.1101/2022.02.13.22270896

**Authors:** Jiwei Li, Jing Wu, Qiuyue Long, Yanan Wu, Xiaoyi Hu, Yukun He, Mingzheng Jiang, Jia Li, Lili Zhao, Shuoqi Yang, Xiaoyong chen, Minghui Wang, Jianshi Zheng, Fangfang Wu, Ruiliang Wu, Lihong Ren, Liang Bu, Houzhao Wang, Ke Li, Lijuan Fu, Guojun Zhang, Yali Zheng, Zhancheng Gao

**Affiliations:** Department of Respiratory, Critical Care and Sleep Medicine, Xiang’an Hospital of Xiamen University, Xiamen, Fujian, China; School of Medicine, Xiamen University, Xiamen, Fujian, China; Department of Clinical Laboratory, Xiang’an Hospital of Xiamen University, Xiamen, Fujian, China; Department of Respiratory and Critical Care Medicine, Peking University People’s Hospital, Beijing, China; Department of Thoracic Surgery, Xiang’an Hospital of Xiamen University, Xiamen, Fujian, China; Department of Pediatrics, Xiang’an Hospital of Xiamen University, Xiamen, Fujian, China; Department of Critical Care Medicine, Xiang’an Hospital of Xiamen University, Xiamen, Fujian, China; Department of Infectious Diseases, Xiang’an Hospital of Xiamen University, Xiamen, Fujian, China; Cancer Research Center and the Department of Breast-Thyroid-Surgery, Xiang’an Hospital of Xiamen University, Xiamen, Fujian, China

## Abstract

The emerging SARS-CoV-2 variants have made great challenges to current vaccine and pandemic control strategies. B.1.1.529 (Omicron), which was classified as a variant of concern (VOC) by the World Health Organization on November 26th, 2021, has quickly become the dominant circulating variant and causing waves of infections. It is urgent to understand the current immune status of the general population given that pre-existing immunity has been established by national vaccination or exposure to past variants. Using sera from 85 individuals (including 21 convalescents of natural infection, 15 cases suffered a breakthrough infection after vaccination, and 49 vaccinated participants without infection history), we showed that the cross-neutralizing activity against VOCs such as Omicron can be detected in 53 (62.4%) cases, although less potent than against the Wuhan-1 strain (WT), with a 3.9-fold reduction in geometric mean neutralizing titer (GMT) (130.7, 95% CI 88.4-193.3 vs 506, 355.8-719.7, respectively). Subgroup analysis revealed significantly enhanced neutralizing activity against WT and VOCs in Delta convalescent sera. The neutralizing antibodies against Omicron were detectable in 75% of convalescents and 44.9% of healthy donors (p = 0.006), with a GMT of 289.5, 180.9-463.3 and 42.6, 31.3-59, respectively. However, the protective effect against VOCs was weaker in young convalescents (aged < 18y), with a detectable rate of 50% and a GMT of 46.4 against Omicron, similar to vaccinees. The pan-sarbecovirus neutralizing activities were not observed in vaccinated SARS-CoV-1 survivors. A booster dose significantly increased the breadth and magnitude of neutralization against WT and VOCs to different degrees than full vaccination. In addition, we showed that COVID-19 inactivated vaccines can elicit Omicron-specific T cell responses. The positive rates of ELISpot reactions were 26.7% (4/15) and 43.8% (7/16) in the full vaccination group and the booster vaccination group, respectively. The neutralizing antibody titers declined while T-cell responses remain robust over 6 months. These findings will inform the optimization of public health vaccination and intervention strategies to protect diverse populations against SARS-CoV-2 variants.

## Introduction

The pandemic of coronavirus disease 2019 (COVID-19) has been ongoing for over two years, making great challenges to the public health system. Numerous genetically distinct lineages with their respective mutations have evolved and have been driving recurrent waves of severe acute respiratory distress syndrome coronavirus-2 (SARS-CoV-2) infection.

The spike glycoprotein of SARS-CoV-2 has two major antigenic domains, the receptor-binding domain (RBD) and the N-terminal domain (NTD), both being the target of neutralizing antibody responses against the virus. Mutations in these regions can lead to increased transmissibility, higher viral binding affinity, and higher antibody escape.^1^ The lineage B.1.1.529 (Omicron) harbors 15 mutations located in the RBD and 8 mutated residues in the NTD relative to the wildtype (WT) virus.^2^ Some mutations on the RBD, including K417N, E484A, and N501Y, are associated with antibody escape.^3^ This heavily mutated strain has raised serious concerns about the diminished protection conferred by pre-existing immunity as soon as it occurred.^4^

Multiple studies have confirmed a large reduction in neutralization titers against Omicron and the failure of many potent monoclonal antibodies (mAbs) to neutralize the variant.^5-7^ The Omicron variant largely evades antibody-mediated immunity and is associated with increased transmissibility, a higher viral load, longer duration of infectiousness, and high rates of breakthrough infection and reinfection, resulting in the Omicron variant rapidly becoming the globally dominant variant.^8,9^ However, the clinical data administered that Omicron-infected individuals have a significantly reduced odds of severe disease compared with individuals infected earlier with the Delta variant.^10^ Some of this reduced severity is probably a result of previous immunity elicited by exposure to past variants, vaccines, and boosters.

The difference between Omicron and other VOCs is that this fifth VOC has emerged at a time when vaccine immunity is increasing in the world. People are “immunologically prepared” after the two-year pandemic of COVID-19. As in China, 87.8% of its population has been vaccinated against SARS-CoV-2. A total of 1.23 billion have received the required two doses to complete vaccinations, adding that 494.4 million had received a booster shot as of January 29, 2022.^11^ However, despite the high national rate, vaccination coverage is still patchy among the elderly and the child and adolescent. Besides, waning antibody titers have raised concerns about the durability of the vaccine.^12^ Furthermore, although current vaccination strategies now propose the administration of a third dose, the clinical efficacy and protection against VOCs remain to be determined. Therefore, it is critical to understand the comprehensive immune responses against SARS-CoV-2 variants in diverse populations.

Here, we characterized the specific humoral and cellular immunity against SARS-CoV-2 variants in different study participants. Using pseudovirus-based neutralization assay, we assessed the cross-reactivity of neutralizing antibodies against the Wuhan-1 (WT), B.1.1.7 (Alpha), B.1.351 (Beta), B.1.617.2 (Delta), and B.1.1.529 (Omicron) variants. Additionally, we evaluated specific T-cell responses against Omicron after a 2- and 3-dose vaccination. We found that WT and VOCs are well neutralized by serum from convalescent individuals who have been vaccinated priorly, although neutralization of Omicron was consistently lower. Booster vaccination enhanced Omicron-specific neutralization, but still at significantly lower levels compared to WT. Robust T cell responses against Omicron were observed in vaccinated healthy donors, especially those who have received a booster dose.

## Methods

### Human subjects

Blood samples were collected from 85 individuals, including 36 delta convalescent patients and 49 vaccinated healthy donors (registered in Chinses Clinical Trial Registry, ChiCTR2100054156). We enrolled convalescents of different age groups to explore possible differences, including the children and adolescents (aged < 18y), the adults (aged 18 - 60y), and the elderlies (aged > 60y). The infection status was confirmed via polymerase chain reactions. The vaccination records were required as well as other demographic data. Serum and peripheral blood mononuclear cells (PBMC) samples were isolated and stored at -80°C until analysis. Briefly, blood from study participants at convalescent time points was processed in a BSL2 laboratory at Xiamen University. Serum samples were heat-inactivated at 56°C for 30 minutes before use. PBMC from all collected blood samples were isolated by Ficoll-Paque density gradient centrifugation. Study approval was obtained from the Ethics Institute of Xiamen University Xiang’an Hospital (XAHLL2021025). All participants provided written informed consent.

### Cell Lines

HEK293T cells were purchased from Procell and cultured in DMEM supplemented with 10% fetal bovine serum (FBS). Cells were grown at 37°C in a 5% CO_2_ setting. Since SARS-CoV-2 uses the SARS-CoV receptor angiotensin-converting enzyme 2 (ACE2) for entry and the transmembrane serine protease 2 (TMPRSS2) for S protein priming, we co-transfected plasmids encoding ACE2 (pLV-ACE2-3xFLAG-IRES-puro, HedgehogBio Science and Technology Ltd.) and TMPRSS2 (pLV-TMPRSS2-GFP, Sino Biological) into 293T cells to generate a stable cell line. The cells were also cultured at 37°C and 5% CO_2_. Confirmation of ACE2 and TMPRSS2 expression in 293T-ACE2-TMPRSS2 cells was done via western blot (Supplementary Figure S1).

### SARS-CoV-2 pseudovirus neutralization assay

The vesicular stomatitis virus (VSV) based pseudoviruses expressing the S protein of several SARS-CoV-2 variants were purchased from Vazyme Biotech, including the WT and 4 VOCs (B.1.1.7 Alpha, B.1.351 Beta, B.1.617.2 Delta, and B.1.1.529 Omicron). The luciferase gene was incorporated into the VSV vector and can be expressed after infection with the pseudotyped virus. The TCID_50_ (50% tissue culture infectious dose) value^13^ was used to quantify virus concentration according to the manufacturer’s instructions.

Neutralization assays were performed on 293T-ACE2-TMPRSS2 cells. Serum samples were 1:16 diluted, followed by a 3-fold serial dilution. The diluted sera (50µL) were mixed with pseudotyped SARS-CoV-2 viruses (650 TCID_50_ per well) in 96-well plates and incubated at 37°C and 5% CO_2_ for 1 h. Then the 293T-ACE2-TMPRSS2 cells were added and co-cultured for the next 24h.^14^ The chemiluminescence signals were measured in relative luminescence units (RLU) using a Bright GloTM luciferase assay system with a GloMax® Navigator Microplate Luminometer. The neutralizing titers (NAT50) were defined as the 50% inhibitory dilution (ID50) which was calculated with the highest dilution of plasma that resulted in a 50% reduction of relative light units compared with virus control. NAT_50_ was calculated with the following formula:

NAT_50_ = 100 × (1 – (value with serum − value in ‘non-infected’) / (value in ‘no serum’ − value in ‘non-infected’).

NAT_50_ below 16 was considered as negative. The NAT_50_ values within groups were summarized as a geometric mean neutralizing titers (GMT) with a 95% confidence interval (95% CI).

### Flow cytometry

The thawed PBMC were incubated overnight at 37°C and 5% CO_2_ in RPMI 1640 + 10% FBS. The next day, cells were harvested and then seeded at 2×10^5^ cells per well in 24-well plates. For either assay, 10 μg/mL recombinant SARS-CoV-2 B.1.1.529 spike RBD protein (Sino Biological) was added to the experimental well while the DMSO in PBS was added to the negative control well. The cells were incubated for 36h at 37°C and 5% CO_2_ before flow cytometry analysis. PBMC were surface stained with fluorescently labeled antibodies to CD3 (FITC), CD19 (PE/Cyanine7), CD4 (PerCP/Cyanine5.5), CD8a (APC), CD56 (BV421), and CD16 (PE) in the dark at 4°C for 30 min. Subsequently, the cells were washed with PBS and stained with Zombie dye (NIR) in the dark at room temperature for 10 min. All FACS antibodies were purchased from Biolegend. After being washed and resuspended, the samples were analyzed using a CytoFlex S cytometer (Beckman Coulter). For each assay, 10,000 events were sampled after the exclusion of debris, doublets, and dead cells. The cellular immune responses against Omicron were measured as a percentage of (CD19^+^) for B or (CD3^+^) for T or (CD16^+^CD56^+^) for NK or (CD3^+^CD4^+^) for CD4^+^ T or (CD3^+^CD8^+^) for CD8^+^ T cells after stimulation of PBMCs with the spike protein.

### Interferon Gamma (IFN-γ) ELISpot assay

IFN-γ secreting T cells were detected by a commercial Human IFN-γ precoated ELISpot kit (Dakewe) according to the manufacturer’s instructions. Briefly, for either assay, approximately 1×10^5^ PBMCs per well were plated into 96-well ELISpot plates, then incubated in the presence of Omicron spike RBD protein (10μg/mL) (experimental wells), phytohemagglutinin (PHA, 5μg/mL) (positive controls), or DMSO (negative controls) for 36h at 37°C and 5% CO2. The cells were subsequently lysed with 4°C deionized water and washed 5 times with PBS. Following the wash, 100μL biotinylated antibody (1:500) and 100μL streptavidin-HRP antibody (1:500) were added to each well and the mixture was incubated at 37°C for 1h. Then, the plate was washed and 100μL/well AEC color developing solution was added. The color reaction was developed for 20 min at room temperature in the dark and stopped by adding 200μL/well of deionized water. Finally, the spot-forming units (SFU), which indicate Omicron-spike-RBD-specific T cells, were counted using an automatic ELISpot Reader. The results were considered positive if experimental wells were no less than twice the negative controls (the signal-to-noise ratio ≥ 2).^15^

### Statistical analysis

Data and statistical analyses were performed using GraphPad Prism 8.0.2 and SPSS 26.0. Flow cytometry data were analyzed using FlowJo 10.4.0. The Pearson χ^2^ test or Fisher’s exact test was performed for a two-group analysis. One-way ANOVA with Tukey’s multiple comparisons test was used to compare differences among multiple groups. Where applicable, the statistical tests used and the definition of the center were indicated in the figure legends. Statistical significance was defined as p < 0.05. Error bars throughout all figures represent 95% confidence interval or one standard deviation where indicated.

## Results

### Population characteristics

Detailed information of the 85 individuals was shown in Table 1. Briefly, the Delta convalescent cases (n = 36) suffered COVID-19 from September 13th to September 18th, 2021 in Xiamen, and the blood samples were obtained between 15 to 40 days post-infection. Among the Delta convalescents, fifteen individuals (15/36, 41.7%) experienced subsequent breakthrough infection after two doses of inactivated virus vaccine (BBIBP-CorV, Sinopharm, Beijing CNBG; or CoronaVac, SinoVac). All the children and adolescents (8/8, 100%) in the Delta convalescent group remain unvaccinated, while nearly half of the adults (10/18, 55.6%) and elderlies (5/10, 50%) received two doses of vaccines. Among the healthy donors, 10 subjects experienced SARS-CoV-1 infection in 2003 in Beijing, who were frontline health care workers during SARS. All the healthy donors received 2 or 3 doses of vaccines except one of the SARS-CoV-1 convalescents. The blood samples were collected between 7 to 381 days after the final vaccination.

**Table 1.**
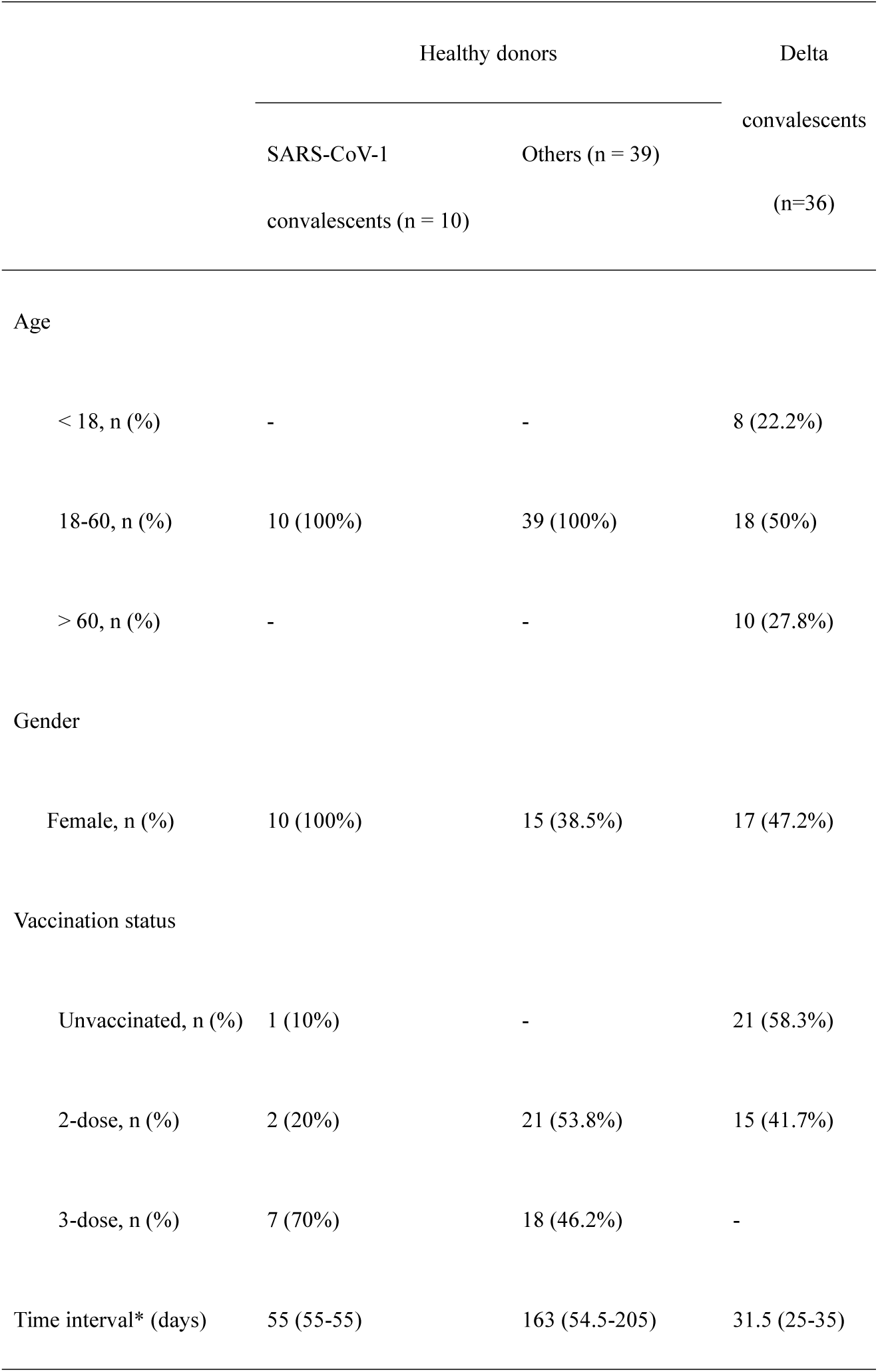
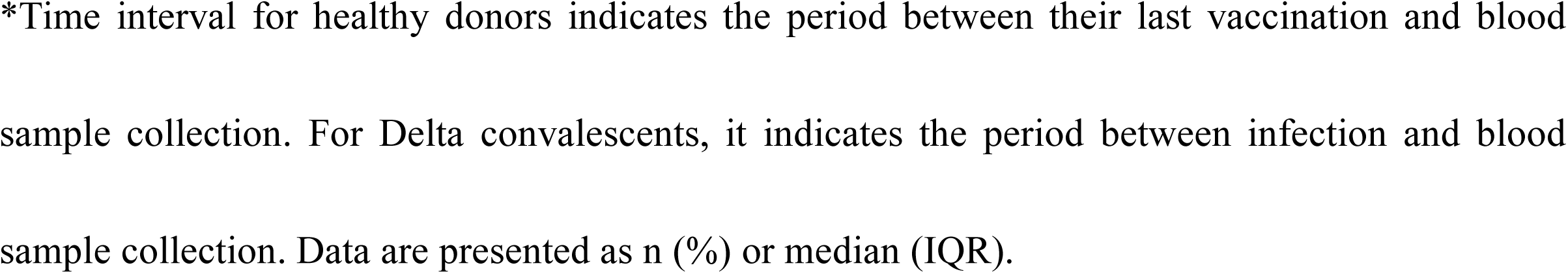
Characteristics of enrolled cases.

|  | Healthy donors |  | Delta |
| --- | --- | --- | --- |
|  | SARS-CoV-1<br>convalescents (n = 10) | Others (n = 39) | convalescents<br>(n=36) |
| Age |  |  |  |
| < 18, n (%) | - | - | 8 (22.2%) |
| 18-60, n (%) | 10 (100%) | 39 (100%) | 18 (50%) |
| > 60, n (%) | - | - | 10 (27.8%) |
| Gender |  |  |  |
| Female, n (%) | 10 (100%) | 15 (38.5%) | 17 (47.2%) |
| Vaccination status |  |  |  |
| Unvaccinated, n (%) | 1 (10%) | - | 21 (58.3%) |
| 2-dose, n (%) | 2 (20%) | 21 (53.8%) | 15 (41.7%) |
| 3-dose, n (%) | 7 (70%) | 18 (46.2%) | - |
| Time interval* (days) | 55 (55-55) | 163 (54.5-205) | 31.5 (25-35) |

\*Time interval for healthy donors indicates the period between their last vaccination and blood sample collection. For Delta convalescents, it indicates the period between infection and blood sample collection. Data are presented as n (%) or median (IQR).

### Reduced cross-neutralizing activity to SARS-CoV-2 variants

As neutralizing antibodies are the major correlate of protection against COVID-19, we first determine the general neutralizing antibody (nAb) responses against WT and four VOCs, including Alpha, Beta, Delta, and Omicron. The results showed a substantial decline in both breadth and potency of all nAbs against the VOCs compared with WT (Figure 1A), no matter the nAbs elicited by vaccination or SARS-CoV-2 infection (Figure 1B). Neutralization against WT was detected in 94.1% (80/85) cases, while in 72.9% (62/85) case, 56.4% (48/85) case, 78.8% (67/85) case and 62.4% (53/85) cases when against Alpha, Beta, Delta, and Omicron variants, respectively (p < 0.0001). The viral neutralization titers (GMT) against the Alpha variant decreased 1.6-fold (a ratio of WT/variant) in pseudovirus assay compared to WT (GMT 506, 95% CI 355.8-719.7), while a 2.7-fold, a 2.2-fold, and a 3.9-fold reduction were observed in GMT against Beta, Delta, and Omicron, respectively. The GMT against the VOCs were 313.5 (95% CI 195.5-502.7), 188.4 (95% CI 115.5-307.1), 232.6 (95% CI 147.6-366.7) and 130.7 (95% CI 88.4-193.3), respectively. These results indicate a general reduction in neutralizing activities of sera against SARS-CoV-2 variants, the degree of decline was in the following order: Omicron > Beta > Delta > Alpha. Generally, the Omicron variant revealed lower neutralizing sensitivity than the Alpha and Delta variants, similar to the Beta variant.

**Figure 1.**
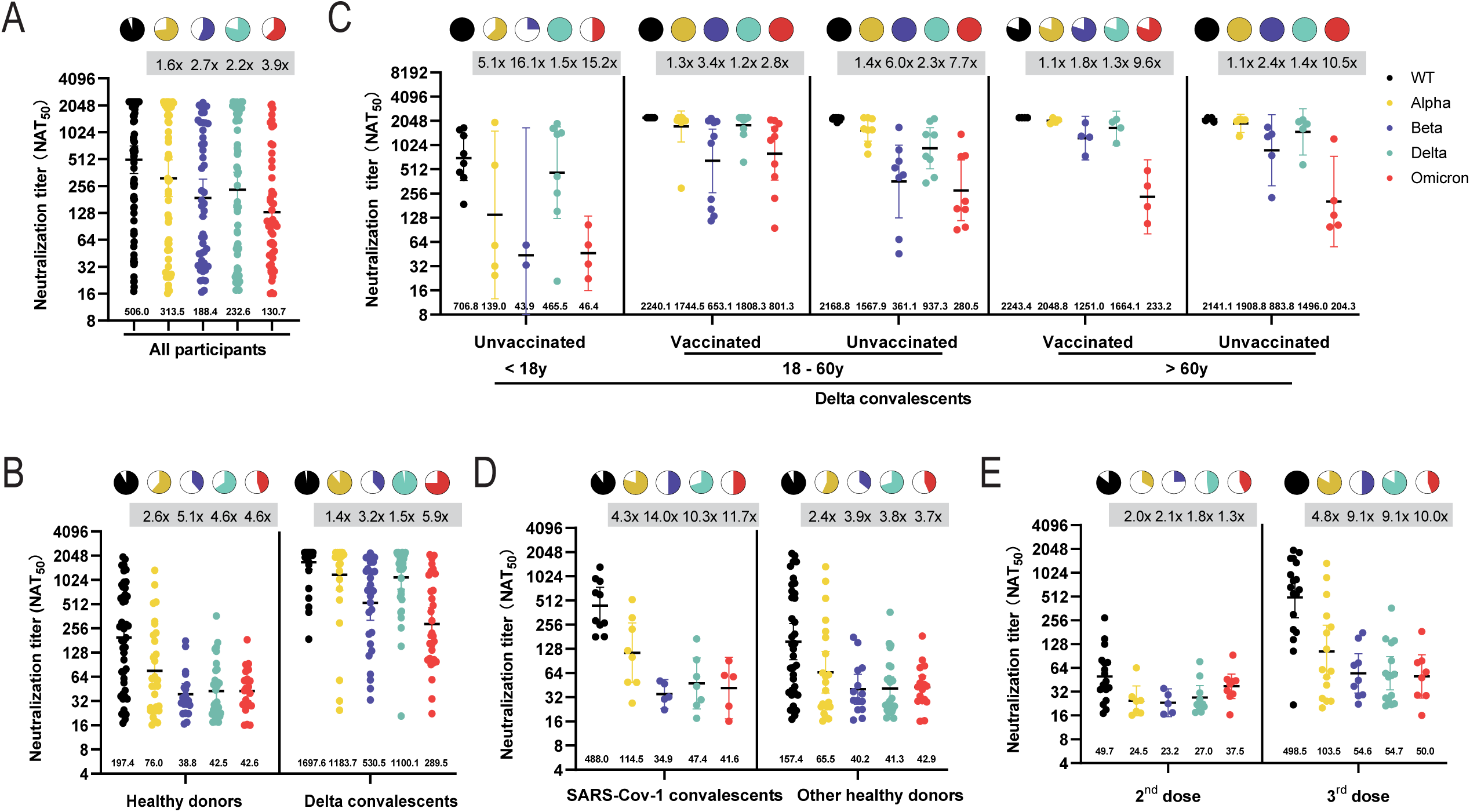
Neutralizing antibody titers against SARS-CoV-2 wild type (WT) and variants of concern (VOCs). (A) The 50% neutralization titers (NAT_50_) were determined via VSV pseudovirus neutralization assay against WT (black dots), Alpha (yellow dots), Beta (purple dots), Delta (green dots), and Omicron (red dots) variants in all samples. (B) Comparison between vaccination sera and convalescent sera. (C) Subgroup analysis of age and vaccination status in Delta convalescents. (D) Subgroup analysis of SARS-CoV-1 infection history in Healthy donors. (E) Subgroup analysis of vaccination status in Healthy donors. Data are presented as scatter dot plots with error bars indicating the geometric mean titers (GMT) with a 95% confidence interval (CI). The GMT values are shown at the bottom of the dots. Fold-change of GMT compared to WT by VOCs are shown at the top of each group. Pie charts show the proportion of vaccinees within each group that had detectable neutralization against the indicated SARS-CoV-2 variants. All neutralization and ELISA assays were conducted in biological duplicates.

### Increased broad-spectrum neutralizing antibodies after Delta infection

To better characterize the neutralization activity of convalescent sera and vaccine sera, we compared the nAbs between the Delta convalescents and the healthy donors (Figure 1B). Impressively, the Delta convalescent sera showed significantly increased neutralizing activities against WT and VOCs. The GMT of convalescent sera against WT and VOCs ranged from 1697.6 to 289.5; while the values ranged from 197.4 to 38.8 in vaccine sera, as shown in Figure 1B and Supplementary Table S1. The nAbs were detectable in 75%-97.2% cases of the Delta convalescents and 38.8%-91.8% participants of the healthy donors.

We further compared the neutralizing activities in different age groups of the Delta convalescents (Figure 1C). We observed a significant increase in the magnitude and breadth of neutralization in the adults and the elderly. However, the increased activities were inapparent in the children and adolescents. For instance, the GMT of convalescent sera against Omicron was 502.6 and 216.7 in the adults and the elderlies, respectively. While the GMT was only 46.4 in the children and adolescents, similar to the vaccinated people (GMT 42.6). Since the children and adolescents had not been vaccinated, we further compared the differences between vaccinated and unvaccinated individuals aged over eighteen. As shown in Figure 1C, people who got infected after being vaccinated (vaccinated-infected) were more resistant to SARS-CoV-2 variants, compared with people who suffered SARS-CoV-2 infection without vaccination (unvaccinated-infected). The neutralizing abilities elicited by infection were less potent in younger cases (aged < 18) when compared with the unvaccinated-infected adults (aged > 18), indicating a focus on the vulnerable population.

### Lack of pan-sarbecovirus neutralizing activity in SARS-CoV-1 survivors

Tan et al. reported a potent cross-clade pan-sarbecovirus nAbs in SARS-CoV-1 convalescents who were immunized with the BNT162b2 mRNA vaccine.^16^ Thus we explored the differences between SARS-CoV-1 convalescents and other healthy donors. All participants received 2 or 3 doses of inactivated vaccines except one of the SARS-CoV-1 convalescents. Although the GMT of the SARS-CoV-1 convalescent sera against WT is 3.1-fold higher than that of the other healthy donors, the potency of neutralization against VOCs was similar between the two subgroups, as shown in Figure 1D. The nAbs against WT or VOCs were undetectable in the only participant who had not been vaccinated.

### A third dose increases the breadth and magnitude of neutralizing antibody responses

A third dose of the COVID-19 vaccines may increase neutralization against VOCs. We applied subgroup analysis between healthy donors who received 2 doses or 3 doses of vaccines (Figure 1E). The booster dose resulted in a 4.8∼10-fold increase in neutralizing activity in 100% participants against WT compared with the second vaccination. We also observed an increase in the breadth and levels of neutralizing antibodies against SARS-CoV-2 variants. Eight out of 18 (44.4%) individuals who received the third dose displayed detectable serum nAbs against Omicron with a GMT of 50 (95% CI 26.6-94.1), compared with 42.9% (9/21) sera detectable with a GMT of 37.5 (95% CI 26.2-53.8) in cases who received the second doses.

We also observed that the nAbs decreased over time after vaccination, as shown in Figure 2A. A sharp decrease in serum nAbs against VOCs and WT was observed at 180-240 days post the final vaccination. The Omicron variant showed a similar decreasing pattern as the Beta and Delta variants, while the Alpha was similar to the WT. NAbs against SARS-CoV-2 variants remain consistent in the Delta convalescents within 30-40 days post-infection (Figure 2B).

**Figure 2.**
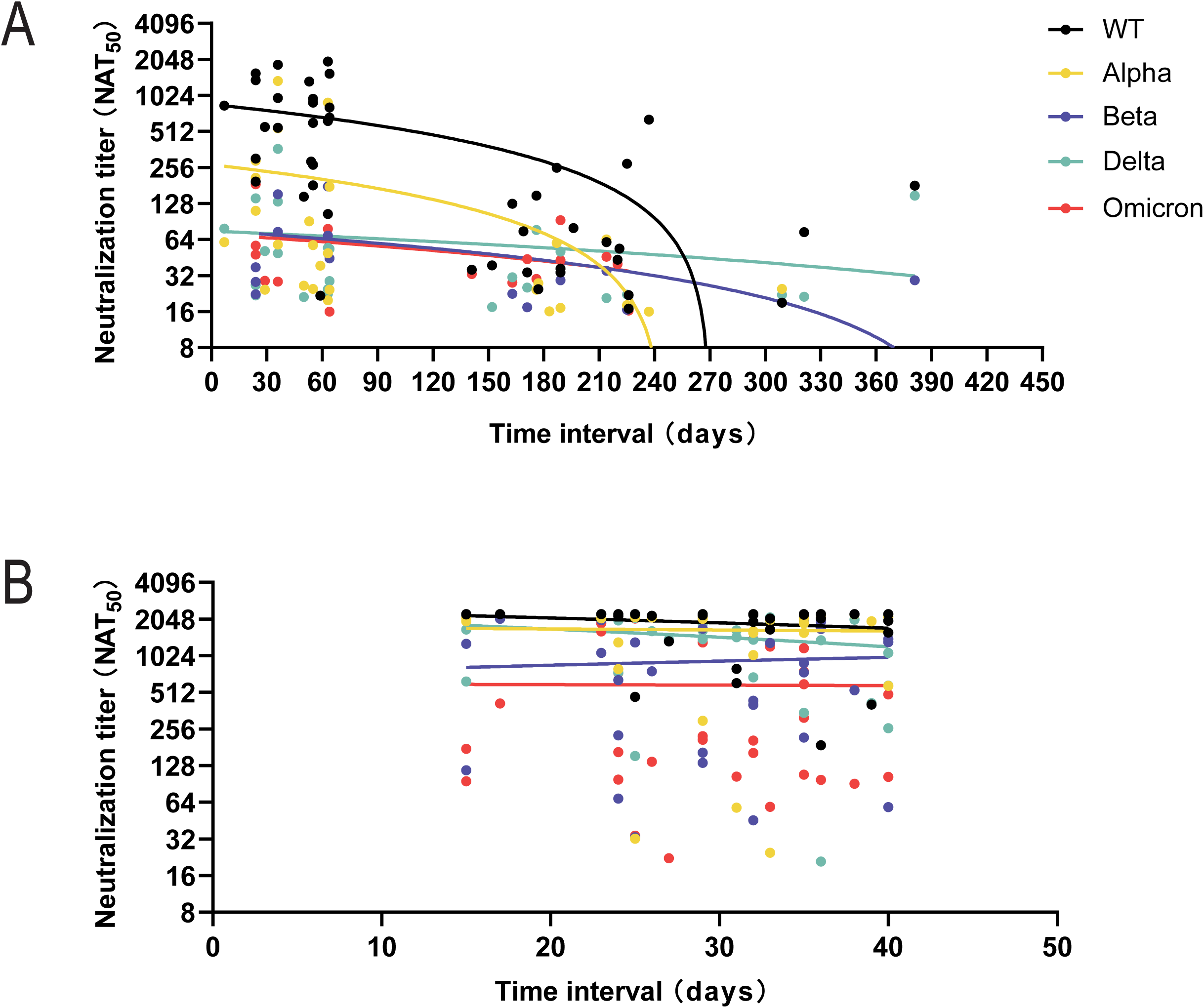
Neutralizing antibody titers decreased over time. The change of nAbs during the interval between last dose vaccination and blood sampling in healthy donors (A) and the time interval between disease onset and blood sampling in Delta convalescents (B). The nAbs show a sharp decline between 180-240 days across all VOCs and WT. In Delta convalescents, the nAbs slightly decreased within 30 days post-infection. The different colored dots represent nAbs against WT (black dots), Alpha (yellow dots), Beta (purple dots), Delta (green dots), and Omicron (red dots).

### Omicron-specific T cell responses in vaccinated healthy donors

Previous studies show that SARS-CoV-2 VOCs partially escape humoral but not T cell responses.^17,18^ The T cell responses could be induced by prior infection, no matter the neutralizing antibody response is reduced or absent.^19^ Here we aimed to elucidate whether the Omicron-specific T cell responses persist before infection, and the association between nAbs and T cell responses.

We first evaluated the PBMC immune responses against Omicron, measured as a percentage of lymphocyte subsets after stimulation of PBMCs with the Omicron-spike protein. As shown in Figure 3A, Supplementary Figure S2, and Supplementary Table S2, we did not detect significant changes in proportions of T, B, and NK cells after stimulation with Omicron-S-RBD protein. Similarly, the subgroup analysis revealed no differences in cellular immune responses in people who received a third dose of vaccination, or who survived from SARS-CoV-1 infection (Figure 3A).

**Figure 3.**
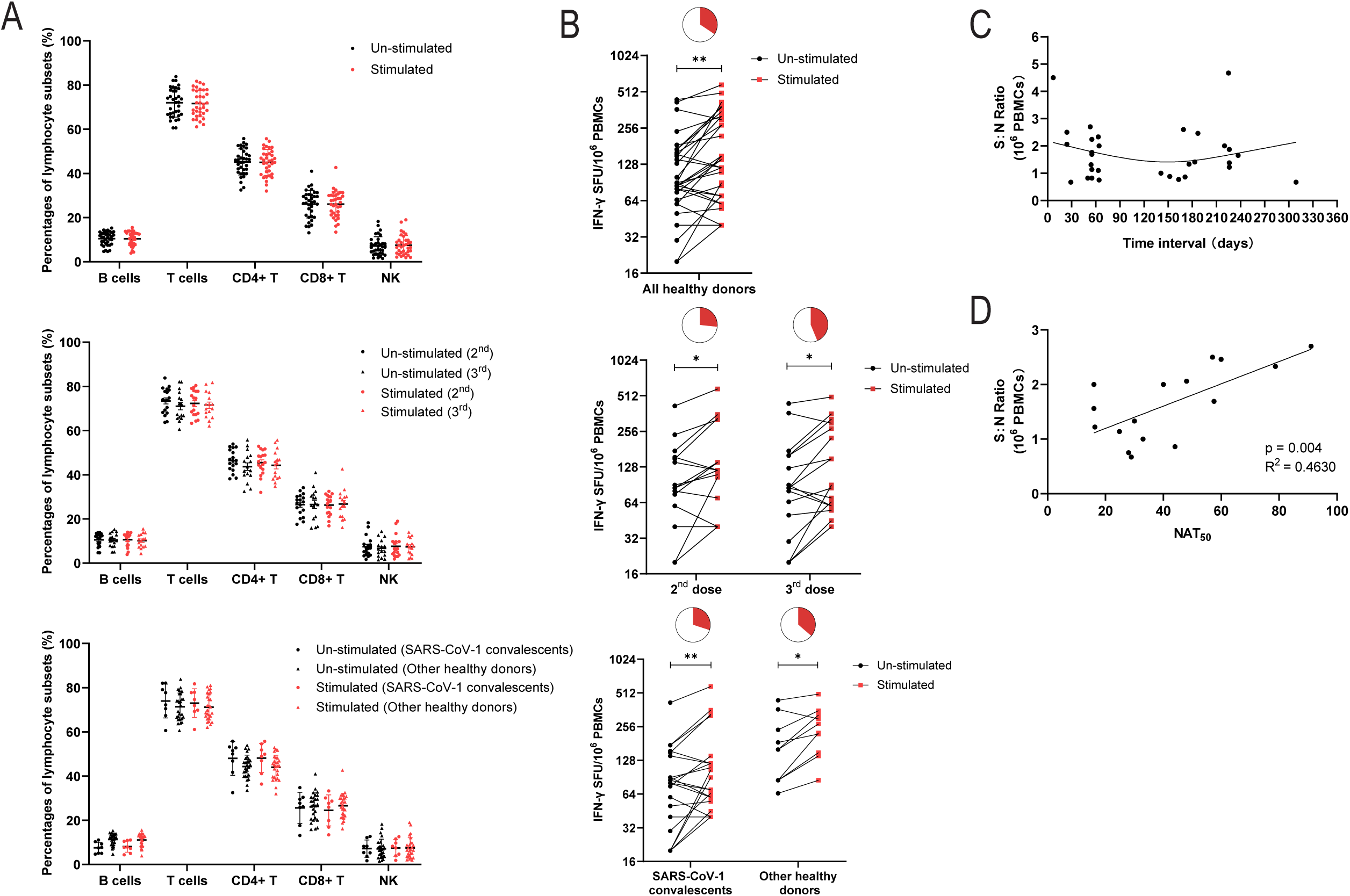
Cellular immune response to recombinant S-RBD proteins of Omicron in healthy donors. (A) The phenotypic analysis results of PBMCs from healthy donors before and after stimulation with Omicron S-RBD protein, followed by subgroup analysis of different vaccination status (having received the 2^nd^ or 3^rd^ dose of vaccination), and then the subgroup analysis between SARS-CoV-1 convalescents and other healthy donors. (B) IFN-γ ELISpot analysis of PBMCs from healthy donors to recombinant Omicron S-RBD proteins, followed by subgroup analysis of different vaccination status (having received the 2^nd^ or 3^rd^ dose of vaccination), and then the subgroup analysis between SARS-CoV-1 convalescents and other healthy donors. The pie charts above represent corresponding proportions of positive ELISpot results within each group. (C) The signal-to-noise (S: N) ratio of SFU at different time intervals after the last dose of vaccine. (D) The correction analysis of signal-to-noise ratio and neutralizing antibody titers (NAT_50_) against Omicron S-RBD protein. * p < 0.05, **p < 0.01.

To further assess virus-specific T cell responses, we performed IFN-γ ELISpot analysis using PBMCs treated with recombinant S-RBD of the Omicron variant (Figure 3B, Supplementary Figure S3, and Supplementary Table S3). We observed that the IFN-γ-secreting S-RBD-specific T cells existed in most participants. Although the numbers of IFN-γ-secreting T cells were similar, the positive reactions (defined as the signal-to-noise ratio ≥ 2) were higher in cases who received a booster dose (7/16, 43.8%), compared to those who received a second dose (4/15, 26.7%), although without significant difference. No significant difference in T cell reaction intensity was detected between SARS-CoV-1 convalescent and other healthy donors, with positive reaction rates of 30% (3/10) and 36.4% (8/22), respectively. Furthermore, the virus-specific immune response of T cells lasted over 6 months post-vaccination (Figure 3C). In addition, a significant positive correlation was observed between the activation intensity of T-cell responses and NAT_50_ (p = 0.004, R^2^ = 0.4630), as shown in Figure 3D. Notably, four participants showed positive T cell reactions while their NAT_50_ values were under the limit of detection. The longest vaccination interval among the four individuals was 225 days, indicating a long-lasting T cell response to SARS-CoV-2, regardless of nAbs levels.

## Discussion

In the current study, we characterized SARS-CoV-2-specific humoral and cellular immunity in different Chinese populations. Sera from vaccinated-infected patients revealed the most powerful cross-neutralizing abilities against SARS-CoV-2 variants. Followed by sera from natural infection convalescents, sera from individuals who received a third dose of inactivated vaccine, and the last, sera from healthy donors who received a second dose of inactivated vaccine. A robust Omicron-specific T cell response was observed in vaccinated healthy donors. We also found that the neutralizing antibody titers were significantly correlated with the intensity of Omicron RBD-specific T-cell responses. The neutralizing antibody titers declined over time while T-cell responses remain consistent or even increase. Since both B and T cells participate in immune-mediated protection to viral infection, our results rationed the current strategy of vaccination, with or without infection history. In addition, the broad antibody responses were weaker in younger convalescents (aged < 18), underscoring an extra focus on these vulnerable populations.

Consistent with most researches, SARS-CoV-2 VOCs showed immune escape capacities from both convalescent sera and vaccine sera. Pseudovirus-based neutralization assay showed reductions in GMTs against the variants compared to the WT, Omicron particularly. However, we observed that breakthrough infection significantly boosts serum neutralizing capacity elicited by post-vaccination. The nAbs against WT and VOCs could be detected in all adult vaccinated convalescents, with approximately 11-fold higher GMT against WT and 18-fold higher against Omicron, compared with vaccinated individuals with no infection history. The GMT against other mutant strains was consistently higher. Recent studies have suggested that vaccination boost cross-variant neutralizing antibodies elicited by SARS-CoV-2 infection.^20,21^ But the effect of breakthrough infection on the neutralizing antibody response is scarce. A preprint observes robust cross-neutralization against Omicron are induced in vaccinees that experienced breakthrough infections.^22^ Furthermore, the comparison between human immune sera following breakthrough infection and vaccination following natural infection showed no difference. They both broadly neutralize SARS-CoV-2 variants to a similar degree.^23^ Together, it is assumed that vaccination plus COVID-19 infection, regardless of which occurs before or after, can boost broad and robust neutralizing antibodies against SARS-CoV-2 variants. The results underscore the importance of vaccination, regardless of infection history.

However, the broad neutralization elicited by infection was much weaker in younger convalescents. The results closely match those obtained in previous studies, which revealed a low protective serological response in both infected and vaccinated adolescents.^24,25^ A reduced breadth of anti-SARS-CoV-2-specific antibodies were observed in children, predominantly generating IgG antibodies specific for the S protein but not the N protein.^25^ These results suggest a distinct humoral immune response in children compared to adults, with implications for age targeting vaccine implementation and effective child protection strategies.

Furthermore, our data suggested a third dose of inactivated vaccine substantially improves neutralization against variants including Omicron. These findings are supported by other studies.^26-29^ However, the variants still showed incomplete escape from booster-enhanced neutralization compared to WT, raising concerns about the efficacy of booster vaccination in the real world. The clinical evidence is still rare. A real-world analysis during the Delta pandemic confirmed that a booster dose substantially lower rates of confirmed Covid-19 and severe illness in adults and the elderly.^30^ Also, a study in nonhuman primates indicates that even low titers of nAbs are sufficient to prevent experimental SARS-CoV-2 infection.^31^ The protection efficacy was apparent if CD8^+^ T cell responses are mounted, indicating that T-cell immune response may ameliorate the deficiency of nAbs in defending against SARS-CoV-2 infection.

The T-cell immune response is an important defense mechanism against the SARS-CoV-2 variants. The virus-specific T cell repertoires could be shaped following natural infection or vaccination.^32-34^ Inspiringly, only 3%-7% of previously identified T cell epitopes are affected by mutations in the various VOCs, indicating minimal escape at the T-cell level.^35,36^ The SARS-CoV-2-specific T cell immune repertoires could still recognize the highly mutated S protein of Omicron.^37^ The study showed approximately 2 in 10^4^ PBMCs were SARS-CoV-2-specific, which is broadly comparable to our findings (1 in 10^4^ PBMCs). Even better, accumulating pieces of evidence suggest that T cell responses to SARS-CoV-2 antigens remain consistent or increase over time, whereas antibody responses wane,^32,33^ consistent with our data. Besides, we observed a higher detection rate of Omicron-specific T cell response in 3-dose vaccinated participants, compared to 2-dose vaccinated individuals. However, whether the difference is caused by the additional antigen exposure from a booster shot or just the result of T cell clonal expansions, remains to be studied.

We further demonstrated the positive correlation between the magnitude of T cell immune response and the titer of sera neutralizing antibodies. Our result is supported by other studies. Zuo et al. demonstrated that the intensity of the T cell response at 6 months correlates both with peak antibody level and a reduced rate of antibody waning against nucleoprotein.^33^ Similarly, Tarke et al. reported a correlation between humoral response and the ability of donors’ PBMCs to become enriched with SARS-CoV-2 spike glycoprotein-reactive CD4^+^ and CD8^+^ T cells.^34^ Together, we assumed that a higher humoral immune response may induce a stronger cellular response. This may further rationalize the current booster vaccination strategy, since the long-term T cell immunological memory to SARS-CoV-2 is essential for the development of herd immunity, which is also the aim of vaccination approaches.

Although analyses were performed in a relatively small number of subjects, we show that breakthrough infection or booster vaccination restored and increased the neutralizing capacity against VOCs including Omicron. Adults and the elderly revealed more effective humoral responses than the child and adolescents. Furthermore, vaccine recipients retain T-cell immunity to the Omicron variant, potentially making up the deficiency of neutralization in preventing or limiting severe COVID-19. The long-term adaptive immune may be key to protection against SARS-CoV-2 variants and even future coronaviruses. Our study supports the current vaccination strategy and calls upon the public health system to prioritize the most vulnerable children.

## Data Availability

The data that support the findings of this study are available within the article and its supplementary materials. Raw data are available from the corresponding authors on reasonable request.

## Acknowledgments

We thank all the participants for their generosity and willingness to engage in the current study. We acknowledge the health care workers (HCWs) of Xiamen University Xiang’an Hospital for their dedication and sacrifice efforts during the COVID-19 local outbreak in Xiamen. We acknowledge the suffering and loss of our COVID-19 patients and that of their families. We acknowledge the HCWs of Peking University People’s Hospital for their donation of blood samples and their serving during the SARS-CoV-1 outbreak.

## Author Contributions

J.W.L., J.W., Y.A.W., Q.Y.L. and X.Y.H. designed the study, conducted the experiments, analyzed and interpreted the results, and wrote the article. Y.K.H., X.Y.H., M.Z.J., and S.Q.Y. did experiments and analyzed and interpreted results. J.L. and L.L.Z. recruited SARS-CoV-1 convalescents and collected the blood samples. X.Y.C., M.H.W., J.S.Z., F.F.W., and R.L.W. recruited SARS-CoV-1 convalescents and collected the blood samples. L.H.R., L.B., H.Z.W., K.L., and L.J.F. coordinated sample acquisition and recruited other healthy donors and convalescents. G.J.Z., Y.L.Z., and Z.C.G. convinced and supervised the study and edited the paper. All authors meet the authorship criteria and approved publication of the manuscript.

## Fund

The study was funded by Xiamen University grant no. 20720200017 and 20720200032.

## References

1. Greaney AJ, Starr TN, Gilchuk P, et al. Complete mapping of mutations to the SARS-CoV-2 spike receptor-binding domain that escape antibody recognition. Cell host & microbe 2021; 29(1): 44–57. e9.

2. McCallum M, Czudnochowski N, Rosen LE, et al. Structural basis of SARS-CoV-2 Omicron immune evasion and receptor engagement. Science (New York, NY) 2022: eabn8652.

3. Harvey WT, Carabelli AM, Jackson B, et al. SARS-CoV-2 variants, spike mutations and immune escape. Nat Rev Microbiol 2021; 19(7): 409–24.

4. Karim SSA, Karim QA. Omicron SARS-CoV-2 variant: a new chapter in the COVID-19 pandemic. Lancet 2021; 398(10317): 2126–8.

5. Dejnirattisai W, Huo J, Zhou D, et al. SARS-CoV-2 Omicron-B. 1.1. 529 leads to widespread escape from neutralizing antibody responses. Cell 2022.

6. Planas D, Saunders N, Maes P, et al. Considerable escape of SARS-CoV-2 Omicron to antibody neutralization. Nature 2021: 1–7.

7. VanBlargan LA, Errico JM, Halfmann PJ, et al. An infectious SARS-CoV-2 B. 1.1. 529 Omicron virus escapes neutralization by therapeutic monoclonal antibodies. Nature medicine 2022: 1–6.

8. Viana R, Moyo S, Amoako DG, et al. Rapid epidemic expansion of the SARS-CoV-2 Omicron variant in southern Africa. Nature 2022: 1–10.

9. Pulliam JR, van Schalkwyk C, Govender N, et al. Increased risk of SARS-CoV-2 reinfection associated with emergence of the Omicron variant in South Africa. MedRxiv 2021.

10. Wolter N, Jassat W, Walaza S, et al. Early assessment of the clinical severity of the SARS-CoV-2 omicron variant in South Africa: a data linkage study. The Lancet 2022.

11. National Health Commission. Available from: http://www.nhc.gov.cn/xcs/fkdt/202201/6b8ac0fcb1f74abe901032877a72c839.shtml. Accessed February 1, 2022.

12. Levin EG, Lustig Y, Cohen C, et al. Waning Immune Humoral Response to BNT162b2 Covid-19 Vaccine over 6 Months. N Engl J Med 2021; 385(24): e84.

13. Nie J, Li Q, Wu J, et al. Quantification of SARS-CoV-2 neutralizing antibody by a pseudotyped virus-based assay. Nat Protoc 2020; 15(11): 3699–715.

14. Di Genova C, Sampson A, Scott S, et al. Production, Titration, Neutralisation, Storage and Lyophilisation of Severe Acute Respiratory Syndrome Coronavirus 2 (SARS-CoV-2) Lentiviral Pseudotypes. Bio Protoc 2021; 11(21): e4236.

15. Moodie Z, Price L, Gouttefangeas C, et al. Response definition criteria for ELISPOT assays revisited. Cancer Immunol Immunother 2010; 59(10): 1489–501.

16. Tan CW, Chia WN, Young BE, et al. Pan-Sarbecovirus Neutralizing Antibodies in BNT162b2-Immunized SARS-CoV-1 Survivors. N Engl J Med 2021; 385(15): 1401–6.

17. Bertoletti A, Tan AT, Le Bert N. The T-cell response to SARS-CoV-2: kinetic and quantitative aspects and the case for their protective role. Oxford Open Immunology 2021; 2(1): iqab006.

18. Geers D, Shamier MC, Bogers S, et al. SARS-CoV-2 variants of concern partially escape humoral but not T cell responses in COVID-19 convalescent donors and vaccine recipients. Science immunology 2021; 6(59): eabj1750.

19. Steiner S, Schwarz T, Corman VM, et al. Reactive T cells in convalescent COVID-19 patients with negative SARS-CoV-2 antibody serology. Frontiers in immunology 2021: 2557.

20. Stamatatos L, Czartoski J, Wan Y-H, et al. mRNA vaccination boosts cross-variant neutralizing antibodies elicited by SARS-CoV-2 infection. Science (New York, NY) 2021; 372(6549): 1413–8.

21. Ibarrondo FJ, Hofmann C, Ali A, Ayoub P, Kohn DB, Yang OO. Previous Infection Combined with Vaccination Produces Neutralizing Antibodies with Potency against SARS-CoV-2 Variants. mBio 2021; 12(6): e02656–21.

22. Miyamoto S, Arashiro T, Adachi Y, et al. Vaccination-infection interval determines cross-neutralization potency to SARS-CoV-2 Omicron after breakthrough infection by other variants. MedRxiv 2022: 2021.12. 28.21268481.

23. Bates TA, McBride SK, Leier HC, et al. Vaccination before or after SARS-CoV-2 infection leads to robust humoral response and antibodies that effectively neutralize variants. Science Immunology 2022: eabn8014.

24. Chen LL, Chua GT, Lu L, et al. Omicron variant susceptibility to neutralizing antibodies induced in children by natural SARS-CoV-2 infection or COVID-19 vaccine. Emerg Microbes Infect 2022: 1–17.

25. Weisberg SP, Connors TJ, Zhu Y, et al. Distinct antibody responses to SARS-CoV-2 in children and adults across the COVID-19 clinical spectrum. Nature immunology 2021; 22(1): 25–31.

26. Ai J, Zhang H, Zhang Y, et al. Omicron variant showed lower neutralizing sensitivity than other SARS-CoV-2 variants to immune sera elicited by vaccines after boost. Emerging microbes & infections 2021; (just-accepted): 1-24.

27. Gruell H, Vanshylla K, Tober-Lau P, et al. mRNA booster immunization elicits potent neutralizing serum activity against the SARS-CoV-2 Omicron variant. Nature medicine 2022: 1–4.

28. Yu X, Wei D, Xu W, et al. Reduced sensitivity of SARS-CoV-2 Omicron variant to antibody neutralization elicited by booster vaccination. Cell Discovery 2022; 8(1): 1–4.

29. Peiris M, Cheng S, Mok CKP, et al. Neutralizing antibody titres to SARS-CoV-2 Omicron variant and wild-type virus in those with past infection or vaccinated or boosted with mRNA BNT162b2 or inactivated CoronaVac vaccines. Research square 2022.

30. Bar-On YM, Goldberg Y, Mandel M, et al. Protection against Covid-19 by BNT162b2 booster across age groups. New England Journal of Medicine 2021; 385(26): 2421–30.

31. Yu J, Tostanoski LH, Peter L, et al. DNA vaccine protection against SARS-CoV-2 in rhesus macaques. Science (New York, NY) 2020; 369(6505): 806–11.

32. Bilich T, Nelde A, Heitmann JS, et al. T cell and antibody kinetics delineate SARS-CoV-2 peptides mediating long-term immune responses in COVID-19 convalescent individuals. Science translational medicine 2021; 13(590): eabf7517.

33. Zuo J, Dowell AC, Pearce H, et al. Robust SARS-CoV-2-specific T cell immunity is maintained at 6 months following primary infection. Nature immunology 2021; 22(5): 620–6.

34. Taborska P, Lastovicka J, Stakheev D, Strizova Z, Bartunkova J, Smrz D. SARS-CoV-2 spike glycoprotein-reactive T cells can be readily expanded from COVID-19 vaccinated donors. Immunity, Inflammation and Disease 2021; 9(4): 1452–67.

35. GeurtsvanKessel CH, Geers D, Schmitz KS, et al. Divergent SARS CoV-2 Omicron-specific T-and B-cell responses in COVID-19 vaccine recipients. MedRxiv 2021.

36. Tarke A, Sidney J, Methot N, et al. Impact of SARS-CoV-2 variants on the total CD4+ and CD8+ T cell reactivity in infected or vaccinated individuals. Cell Reports Medicine 2021; 2(7): 100355.

37. Ahmed SF, Quadeer AA, McKay MR. SARS-CoV-2 T Cell Responses Elicited by COVID-19 Vaccines or Infection Are Expected to Remain Robust against Omicron. Viruses 2022; 14(1).

