## Supplementary Figure Legend for "Comprehensive humoral and cellular immune responses to SARS-CoV-2 variants in diverse Chinese populations: A benefit perspective of national vaccination"

**Supplementary Figure S1. Western blot analysis confirmed the expression of ACE2 and TMPRSS2 in 293T-ACE2-TMPRSS2 cells.**

Plasmids encoding ACE2 and TMPRSS2 were co-transfected into 293T cells to generate a stable cell line (293T-ACE2-TMPRSS2 cells). β-actin was used as the internal control.

**Supplementary Figure S2. The gating strategy of Flow cytometry.**

Representative gating of $CD19^+$ B cells, $CD3^+$ T cells, $CD4^+$ T cells, $CD8^+$ T cells and $CD16^+CD56^+$ NK cells from donor PBMCs is shown. Briefly, live cells are gated as Zombie UV-. Cells were then gated as $CD19^+$, $CD3^+$ or $CD19^-CD3^-$ cells. T cells were further subdivided into either $CD8^+$ or $CD4^+$ populations. $CD3^-CD19^-$ cells were then divided into $CD56^+CD16^+$ NK cells. For each assay, 10,000 events were sampled after the exclusion of debris, doublets, and dead cells.

**Supplementary Figure S3. Representative ELISpot results.**

Representatively positive ELISpot results from different healthy donors. Approximately $1\times10^5$ PBMCs per well were incubated in the presence of Omicron spike RBD protein (10μg/mL) (experimental wells), phytohemagglutinin (PHA, 5μg/mL) (positive controls), or DMSO (negative controls) for 36h at 37℃ and 5% $CO_2$. The spot-forming units (SFU) were counted using an automatic ELISpot Reader and the numbers of SFU were shown at the top right of the figure.
