## Supplementary figures and images for "Comprehensive humoral and cellular immune responses to SARS-CoV-2 variants in diverse Chinese populations: A benefit perspective of national vaccination"

### Supplementary Figure S1

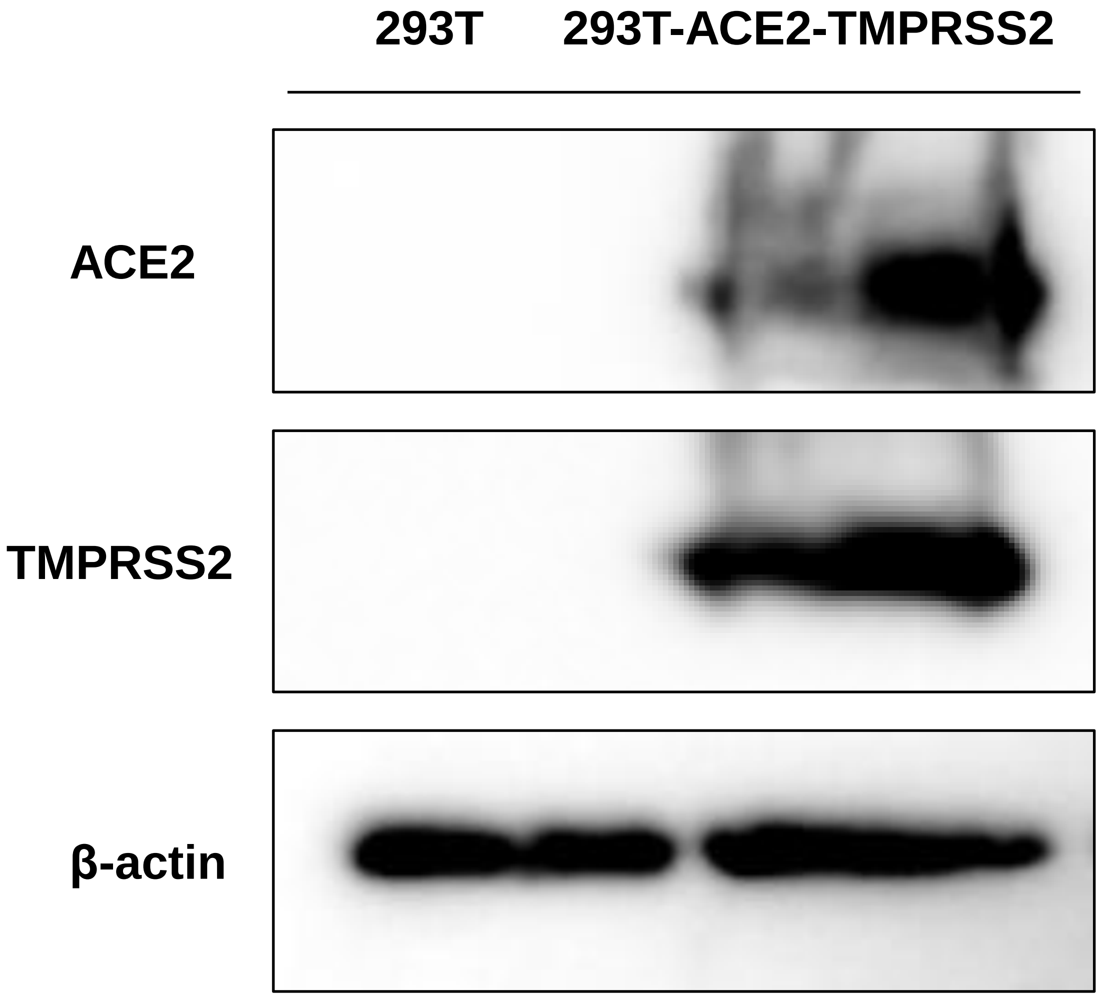

### Supplementary Figure S2

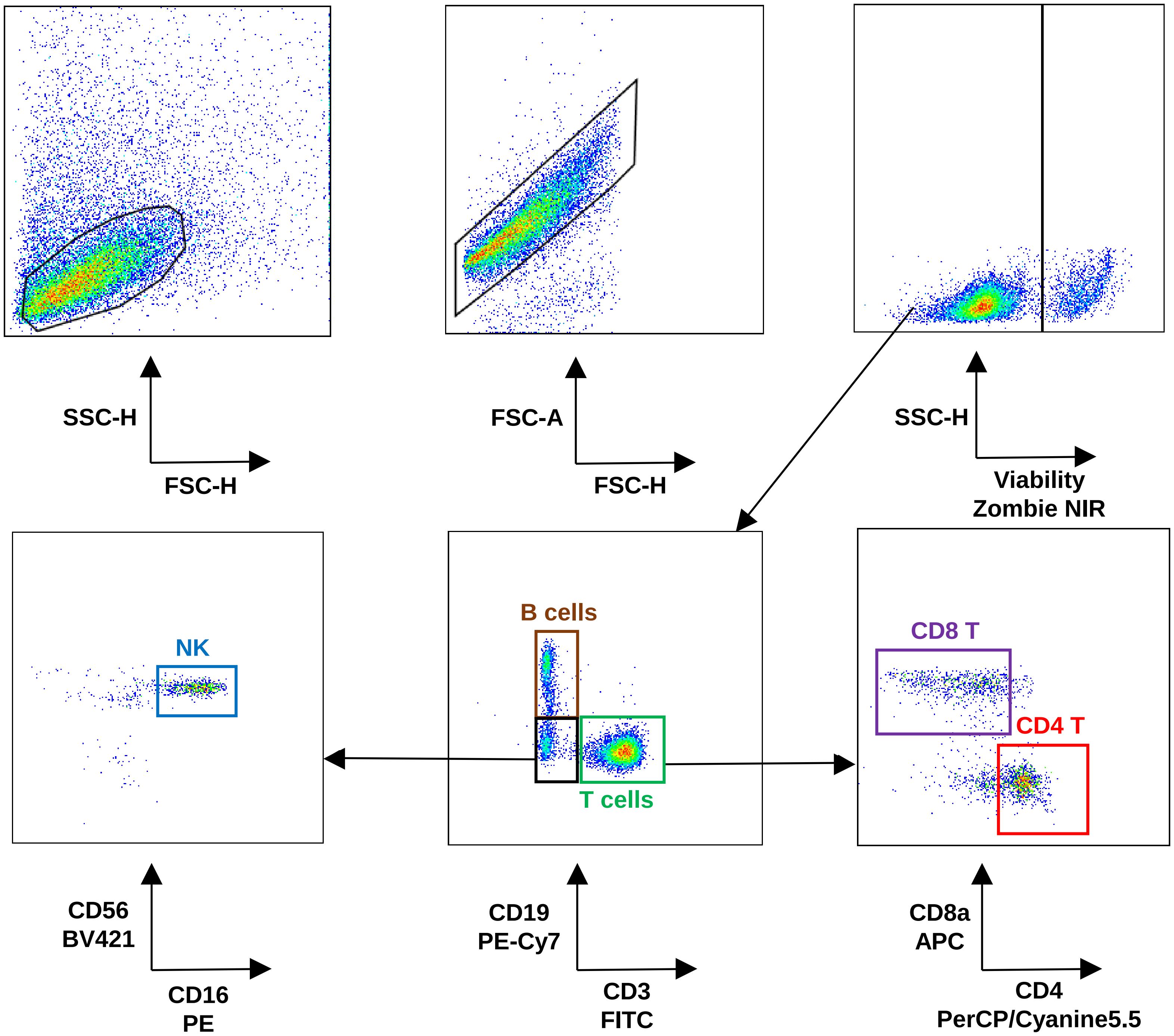

### Supplementary Figure S3

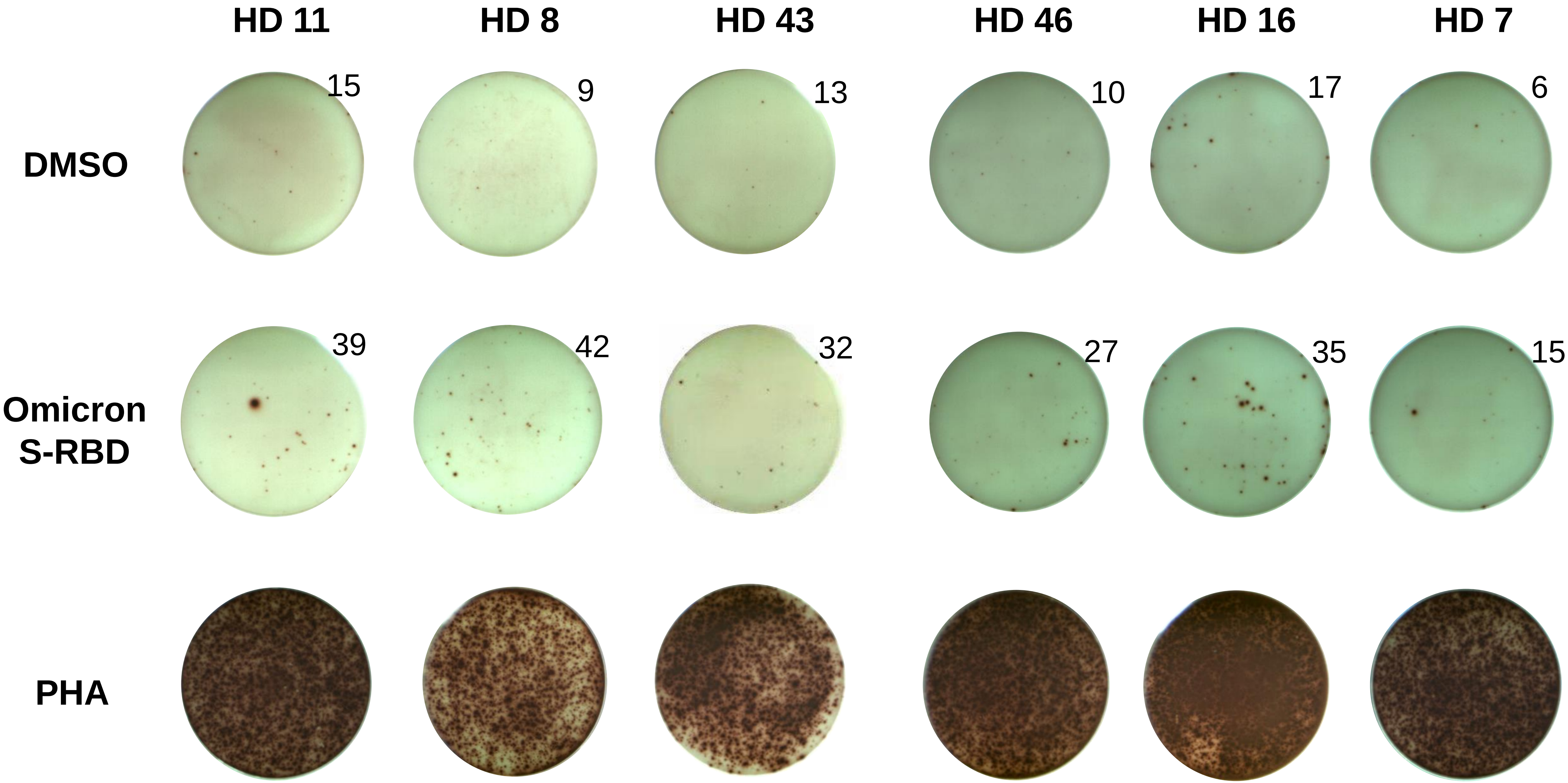
