## Supplementary Tables for "Comprehensive humoral and cellular immune responses to SARS-CoV-2 variants in diverse Chinese populations: A benefit perspective of national vaccination"

**Supplementary Table S1. GMT values with 95%CI in different subgroups**

| | | **Delta convalescents** (n = 36) | | | | | | | | **Healthy donors** (n = 49) | | | | |
|---|---|---|---|---|---|---|---|---|---|---|---|---|---|---|
| | **All participants** (n = 85 ) | All convalescents | <18y | 18-60y (n = 18) | | | >60y (n=10) | | | All healthy donors (n = 49) | SARS-CoV-1 convalescents (n = 10) | Other Healthy donors (n = 39) | | |
| | | | unvaccinated (n = 8) | All 18-60y (n = 18) | Vaccinated (n = 10) | Unvaccinated (n=8) | All > 60y (n = 10) | Vaccinated (n = 5) | Unvaccinated (n = 5) | | | All | $2^{nd}$ dose (n = 21) | $3^{rd}$ dose (n = 18) |
| WT | 506 (355.8-719.7) | 1697.6 (1383-2083) | 706.8 (373.8-1337) | 2208.2 (2168-2250) | 2240.1 (2231-2250) | 2168.8 (2077-2265) | 2186 (2110-2264) | 2243.4 (2240-2246) | 2141.1 (2001-2291) | 197.4 (93.9-263.8) | 488 (283.7-839.5) | 157.4 (93.9-263.8) | 49.7 (34.3-72) | 498.5 (275.3-902.8) |
| Alpha | 313.5 (195.5-502.7) | 1183.7 (759.2-1846) | 139 (12.6-1533) | 1663.7 (1287-2151) | 1744.5 (1118-2722) | 1567.9 (1143-2150) | 1969.8 (1743-2226) | 2048.8 (1823-2302) | 1908.8 (1472-2475) | 76 (36.1-118.8) | 114.5 (48.7-269.3) | 65.5 (36.1-118.8) | 24.5 (15.9-37.9) | 103.5 (48.2-222.5) |
| Beta | 188.4 (115.5-307.1) | 530.5 (324.2-868.1) | 43.9 (1.1-1678) | 501.9 (268.4-938.2) | 653.1 (262.9-1622) | 361.1 (127.9-1020) | 1031.4 (625.1-1702) | 1251 (670.6-2334) | 883.8 (320.6-2436) | 38.8 (26.2-61.8) | 34.9 (23-53.1) | 40.2 (26.2-61.8) | 23.2 (15.5-34.7) | 54.6 (30.9-96.7) |
| Delta | 232.6 (147.6-366.7) | 1100.1 (782.6-1546) | 465.5 (125.1-1732) | 1350.3 (984.6-1852) | 1808.3 (1353-2417) | 937.3 (520.8-1687) | 1568.5 (1130-2177) | 1664.1 (1022-2709) | 1496 (769.3-2909) | 42.5 (29.6-57.6) | 47.4 (22.9-53.1) | 41.3 (29.6-57.6) | 27 (19.1-38.1) | 54.7 (33.9-88.5) |
| Omicron | 130.7 (88.4-193.3) | 289.5 (180.9-463.3) | 46.4 (16-134.7) | 502.6 (283.3-891.7) | 801.3 (374.9-1713) | 280.5 (117.7-668.8) | 216.7 (113.2-414.9) | 233.2 (80.8-672.6) | 204.3 (56-745.2) | 42.6 (31.3-59) | 41.6 (17.2-100.8) | 42.9 (31.3-59) | 37.5 (26.2-53.8) | 50 (26.6-94.1) |

**Supplementary Table S2. Proportions of lymphocyte subsets in PBMCs from vaccines before and after stimulation.**

| | Un-stimulated | | | | | Stimulated | | | | |
|---|---|---|---|---|---|---|---|---|---|---|
| | Total B (%) | Total T (%) | $CD4^+T$ (%) | $CD8^+T$ (%) | NK (%) | Total B (%) | Total T (%) | $CD4^+T$ (%) | $CD8^+T$ (%) | NK (%) |
| Healthy donors (n = 35) | 10.5 ± 3 | 72±6.5 | 45.2± 5.9 | 26.1± 6.3 | 7.1 ± 4.2 | 10.4 ± 3.1 | 71.6 ± 6 | 45 ± 5.9 | 26.1 ± 5.9 | 7.5 ± 4.4 |
| SARS-CoV-1 convalescents (n = 8) | 7.6 ± 2.5 | 74 ± 7.6 | 48 ± 7.6 | 25.6 ± 7.1 | 7.2 ± 3.9 | 8.1 ± 2.6 | 73 ± 6.5 | 48.2 ± 6.6 | 24.5 ± 7 | 7.5 ± 3.8 |
| Other healthy donors (n = 27) | 11.4 ± 2.6 | 71.4 ± 6.1 | 44.4 ± 5.1 | 26.3 ± 6.2 | 7.1 ± 4.4 | 11.1 ± 2.9 | 71.2 ± 5.9 | 44.1 ± 5.4 | 26.6 ± 5.5 | 7.5 ± 4.6 |
| $2^{nd}$ dose (n = 18) | 10.5 ± 3.3 | 73.4 ± 5.9 | 46.5 ± 5 | 26.5 ± 4.9 | 7.4 ± 4.6 | 10.6 ± 3.2 | 72.3 ± 5.9 | 45.5 ± 5.5 | 26.3 ± 4.7 | 7.6 ± 5 |
| $3^{rd}$ dose (n = 16) | 10.4 ± 3.2 | 71.1 ± 6.6 | 43.7 ± 6.8 | 26.5 ± 7.1 | 6.7 ± 4 | 10.2 ± 3.1 | 71.5 ± 5.8 | 44.3 ± 6.5 | 26.8 ± 6.5 | 7.4 ± 3.8 |

Data were presented as mean ± SD. Only one of the SARS-CoV-1 convalescents remained unvaccinated.

**Supplementary Table S3. Descriptive statistics of SFUs per $10^6$ PBMCs in healthy donors in ELISpot experiment.**

| | Un-stimulated | Stimulated |
|---|---|---|
| Healthy donors (n = 32) | 125.9 ± 105.9 | 188.1 ± 147.9 |
| SARS-CoV-1 convalescents (n = 10) | 177 ± 124.8 | 256 ± 119.2 |
| Other healthy donors (n = 22) | 102.7 ± 89.9 | 157.3 ± 151.7 |
| 2nd dose (n = 15) | 124.7 ± 98 | 198 ± 166 |
| 3rd dose (n = 16) | 123.8 ± 118.4 | 177.5 ± 138.6 |

Data were presented as mean ± SD.
